# Predictors of characteristics associated with negative first SARS-CoV-2 PCR testing despite final diagnosis of COVID-19, and association with treatment and outcomes. The COVID-19 RT-PCR Study

**DOI:** 10.1101/2020.09.14.20194001

**Authors:** Jean-Baptiste Lascarrou, Gwenhael Colin, Aurélie Le Thuaut, Nicolas Serck, Mickael Ohana, Bertrand Sauneuf, Guillaume Geri, Jean-Baptiste Mesland, Gaetane Ribeyre, Claire Hussenet, Anne Sophie Boureau, Thomas Gille

## Abstract

**Background:** Reverse transcriptase-polymerase chain reaction (RT-PCR) testing is an important tool for the diagnosis of coronavirus disease 2019 (COVID-19). However, performance concerns have recently emerged, especially about its sensitivity.. We hypothesized that clinical, biological and radiological characteristics of patients with false negative first RT-PCR testing, despite final diagnosis of COVID-19, might differ from patients with positive first RT-PCR.

**Methods:** Case – control, multicenter study in which COVID-19 patients with negative first RT-PCR testing were matched to patients with positive first RT-PCR on age, gender and initial admission unit (ward or intensive care).

**Results:** Between March 30, and June 22, 2020, 80 cases and 80 controls were included. Neither proportion of death at hospital discharge, nor duration of hospital length stay differed between “case” and “control” patients (P = 0.80 and P = 0.54, respectively). In multivariate analysis, headache (adjusted OR: 0.07 [0.01; 0.49]; P = 0.007) and fatigue/malaise (aOR: 0.16 [0.03; 0.81]; P = 0.027) were associated with lower risk of false negative, whereas platelets > 207.10^3^.mm^−3^ (aOR: 3.81 [1.10; 13.16]; P = 0.034) and CRP > 79.8 mg.L^−1^ (aOR: 4.00 [1.21; 13.19]; P = 0.023) were associated with higher risk of false negative.

**Interpretation:** Patients with suspected COVID-19 and higher inflammatory biological signs expected higher risk of false negative RT-PCR testing. Strategy of serial RT-PCR testings must be rigorously evaluated before adoption by clinicians.

## INTRODUCTION

Since December 2019, a novel coronavirus named severe acute respiratory syndrome coronavirus 2 (SARS-CoV-2) emerged in Wuhan city in China, and rapidly spread throughout China, Asia and worldwide (1). On September 1, 2020 more than 25 000 000 patients had been infected and 850 000 had died from Coronavirus disease 2019 (COVID-19).

Coronaviruses are enveloped RNA viruses that are broadly distributed among humans, other mammals and birds, causing respiratory, enteric, hepatic, and neurologic diseases. Identification and sequencing of SARS-CoV-2 was achieved by a Chinese team with rapid communication of their results, allowing clinicians worldwide to perform reverse transcriptase polymerase chain reaction (RT-PCR) testing in suspected patients (2). Increasing literature has emerged to highlight multiple presentations of COVID-19, even though respiratory symptoms are predominant (3). Given the presence of multiple, nonspecific symptoms, accurate diagnosis is the cornerstone of health care, with possible implications for isolation, corticosteroids administration and location of hospitalization (ward / intensive care unit).

Recently, performance concerns arose about RT-PCR testing, especially for sensitivity, as highlighted by the report of 2 false negative COVID-19 RT-PCR testings by Li *et al*. (4). In a cohort of 219 confirmed COVID-19 patients matched to 205 patients with viral pneumonia from other origin, Bai *et al*. (5) found than chest computed tomography (CT) scan outperformed nasopharyngeal testing to rule in or rule out COVID-19 disease. While the analytic performance of SARS-CoV-2 RT-PCR testings is well described (6), clinical performance can be diminished by several factors: low levels of shedding (7), site of sample collection (8) and technical background of nurses and technicians in charge. Whereas symptoms of COVID-19 infection are not specific (fever, cough, fatigue and lymphopenia) (1), RT-PCR testing and result interpretation can be a concern for clinicians.

We hypothesized that clinical and/or biological and/or radiological characteristics of patients with false negative first SARS-CoV-2 RT-PCR testing despite final diagnosis of COVID-19 could be different from patients with positive first SARS-CoV-2 RT-PCR; and that patients with false negative first SARS-CoV-2 RT-PCR may expect better outcome than patients with positive first SARS-CoV-2 RT-PCR. To answer this, we performed a case-control study in which COVID-19 patients with negative first SARS-CoV-2 RT-PCR testing were matched to patients with positive first SARS-CoV-2 RT-PCR.

## METHODS

### Design and definition of case and control

Elevenhospitals from France and Belgium participated to this multicenter, retrospective, case-control analysis. Cases were defined as patients admitted in hospital with final diagnosis of COVID-19 despite a negative first SARS-CoV-2 RT-PCR testing. Controls were patients from the same hospital, matched on gender, age and initial admission unit (ward or intensive care) with positive first SARS-CoV-2 RT-PCR testing.

### Eligibility

Inclusion criteria were:

–Age > 18 years.
–Hospitalization for infectious condition and final diagnosis of COVID-19:

- With negative first SARS-CoV-2 RT-PCR for cases.
- With positive first SARS-CoV-2 RT-PCR for controls.

Non-inclusion criteria were:

–Biological identification of another virus responsible for the pneumonia.
–Pregnancy, recent delivery or lactation.
–Adult under guardianship or curatorship.

### Outcomes

Our primary objective was to identify factors associated with higher risk of false negative first SARS-CoV-2 RT-PCR testing (regarding sensitivity of RT-PCR testing). Secondary outcomes were: delivered treatments, need for mechanical ventilation, duration of mechanical ventilation, occurrence of acute respiratory distress syndrome (ARDS) and outcome at hospital discharge.

### Collected data

All data in the eCase Report Form (eCRF) were anonymized, and no data could be traced back to the patient’s identity. Each local investigator filled an eCRF to collect data (Castor EDC, Amsterdam, Netherlands). Collected data were: matching characteristics (age, gender, location); baseline demographics (comorbidities); clinical and biological characteristics at hospital admission; history of symptoms; radiological findings; RT-PCR testing results (first RT-PCR testing ± final RT-PCR testing if positive for “case” patients); other pathogen testings and results; antiviral treatments; outcomes; final diagnostic modalities for “case” patients.

### Ethics

The study was approved by the appropriate ethics committees (For France: *Comité d’éthique de la Société de Réanimation de Langue Française*, #20–26; and for Belgium: *Comité d’Ethique 045 Clinique Saint Pierre)* which waived consent according to collected data.

### Statistical analysis

Statistical analysis were performed according to STROBE guidelines (9). Cases were matched with controls from the same hospital on age, gender, and initial admission unit (ward or intensive care) on 1:1 basis. Qualitative variables were described as numbers (%) and quantitative variables as means ± SD if normally distributed or medians [25^th^ – 75^th^ percentiles] otherwise. Mortality and hospitalization rate were compared between cases and controls using conditional logistic regression to take into account paired data. Conditional logistic regression models were used to identify factors associated with negative RT-PCR test. Step by step backward selection was applied. Predefined factors associated with negative RT-PCR testing at *P* values ≤ 0.2 by univariate analysis were then introduced in a multiple logistic regression model with retaining of variable associated with P value ≤ 0.1 (conservative approach). Homesher-Lemeshow test and visual inspection of residues were used to ensure quality of the regression. Quantitative variables were dichotomized according to their median. Selection of collinear variables was performed according to their clinical relevance. Model selection was based on Akaike information criterion (AIC) (10). Regarding the importance of duration between symptom onset and RT-PCR testing in previous literature, this variable was forced in all models. All statistical analyses were performed using SAS (Microsoft, Redmond, CA, USA).

### Sample size

Regarding exploratory nature of our study, we did not set sample size but we targeted at least 50 patients and 50 controls (100 patients).

## RESULTS

Between March 30, and June 22, 2020, 82 “case” patients were identified. Due to the non-inclusion of 2 related matched controls, 2 “case” patients were excluded from analysis. At last, 80 cases and 80 controls were finally included. Patients were mainly males (66.3%), with a mean age of 64.1 ± 16.8 years old, and more frequently admitted in ward (71.3%).

Among the 80 included cases, a chest radiography was performed for 25 patients (main finding: normal (N = 1), ground-glass opacities (N = 4), local patchy opacities (N = 1), bilateral patchy opacities (N = 12), interstitial abnormalities (N = 7)) and a chest CT scan was performed for 75 cases (main finding: normal (N = 1), ground-glass opacities (N = 69), interstitial abnormalities (N = 4); not described (N = 1)).

RT-PCR testing was realized 6 [2.5–10.5] days after symptom onset for cases and 5 [1.0–9.0] days for controls (P = 0.27). For 11 cases with subsequent positive RT-PCR testing, it was performed 11.0 [9.0–16.0] after symptom onset.

Final diagnosis of COVID-19 cases (N = 80) were established on (multiple answers were allowed for each patient): subsequent oropharyngeal positive RT-PCR (N = 9), subsequent expectoration positive RT-PCR (N = 1), subsequent tracheal positive RT-PCR (N = 4), chest CT scan (N = 71), contagion from closed family member (N = 13), returned from infected cluster (N = 2), serology (N = 2).

Clinical participant characteristics for cases and controls are detailed in Table 1, and their biological characteristics in Table 2.

On univariate analysis, fatigue/malaise (P = 0.048), headache (P = 0.048), history of fever (P = 0.020), myalgia (0.024) and elevation of hepatic enzymes (alanine aminotransferase (ALAT) aspartate aminotransferase (ASAT), P = 0.024 for both) were associated with lower risk of first negative RT-PCR testing (OR < 1); whereas platelets upper than 207 per 10^3^.mm^−3^ (P = 0.002), white blood cells > 6.95 per 10^3^.mm^−3^ (P = 0.0003) and C-reactive protein (CRP) > 79.8 mg.L^−1^ (P = 0.28) were associated with higher risk of first negative RT-PCR testing (OR > 1).

Because ASAT and ALAT were collinear of platelet count and white blood cells were collinear of CRP, they were not included in multivariate analysis. The result of multivariate analysis is depicted on Figure 1 with an AIC: 54.8 and BIC: 69.1.

**Figure 1:**
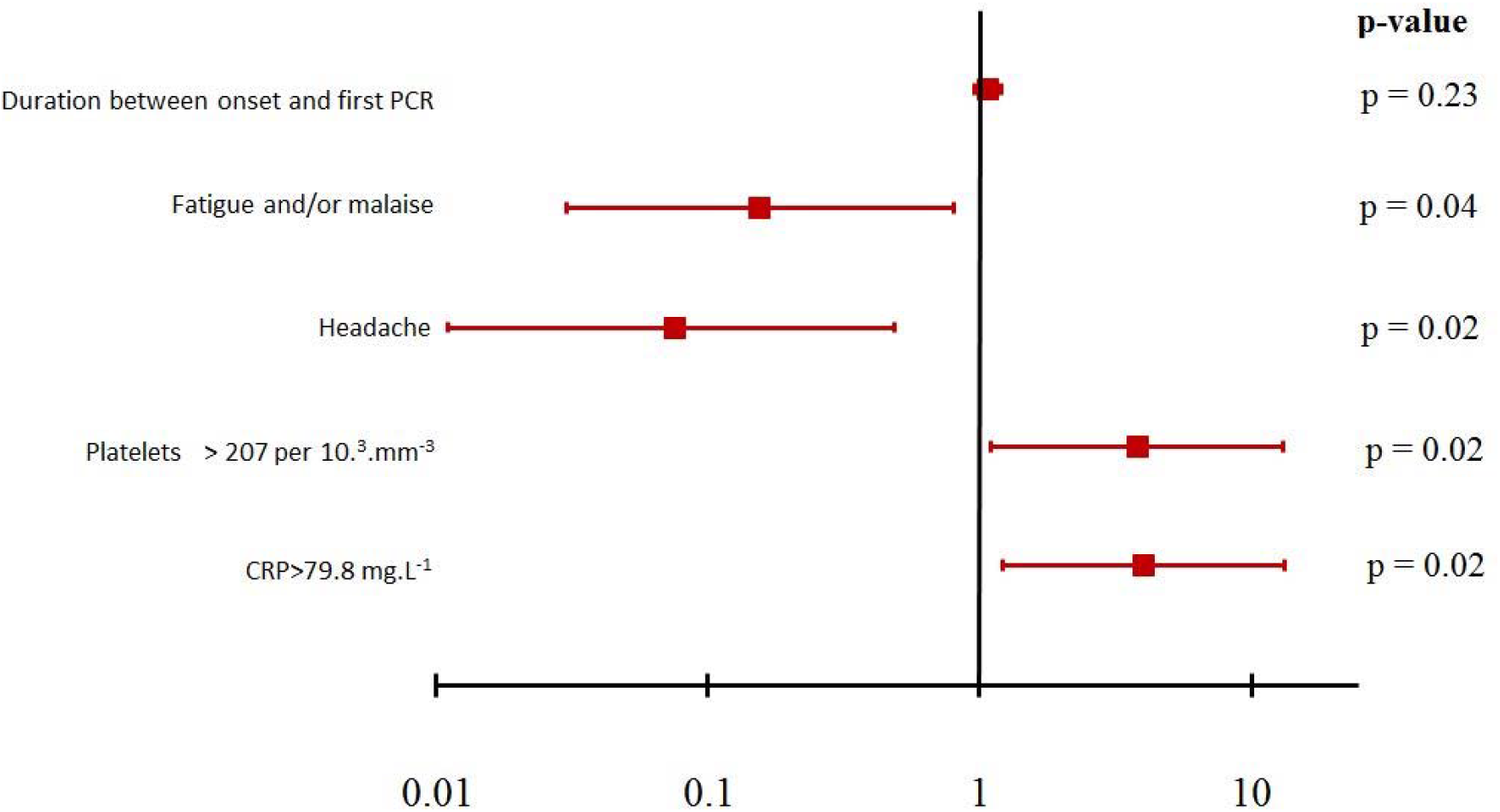
Forrest plot of multivariable analysis of factors associated with first false negative SARSCoV-2 RT-PCR testing

The proportion of “case” and “control” patients who received at least one treatment (chloroquine, corticosteroids, lopinavir/ritonavir, macrolids or tocilizumab) did not differ (P = 0.26) (Table 3). Mechanical ventilation was required for 10 (12.7%) cases and 14 (17.7%) controls (P = 0.177), for a duration of 21 [16–35] days for cases and 15 [5–21] days for controls (P = 0.197).

Neither proportion of death at hospital discharge, nor duration of hospital length stay differed between “case” and “control” patients (P = 0.80 and P = 0.54, respectively).

## DISCUSSION

In this study, patients with higher CRP elevation or higher platelet count expected higher risk of false negative SARS-CoV-2 RT-PCR testing, whereas patients with non-specific symptoms such as headache or fatigue/malaise expected lower risk of false negative RT-PCR. Duration between symptom onset and time of RT-PCR testing was not associated with false negative result.. Finally, patients with false negative testing did not receive different treatments and did not expect different outcome either, according to the proportion of patients requiring mechanical ventilation and to mortality at hospital discharge.

Tree main conclusions can be drawn. First, duration between symptom onset and time to RTPCR testing was not associated with positivity. Such result can probably be explained in part by difficulties in medical history examination, especially in older patients,like what was previously described for other diseases (11), as mean age in our cohort was 64 ± 17 years old. Additionally, frequent presence of delirium (up to 25%) in geriatric patients can reduce the reliability of reported symptom duration (12).

Second, the association between high CRP level and higher risk of negative RT-PCR testing is of interest, because it is an argument for mortality associated with cytokine storm, regardless of viral load (13). It is a matter of concern, given the results of RECOVERY trial, in which corticosteroids were the only treatment proven to be effective to reduce mortality for patients with COVID-19 (14). Small proportion of our patients received corticosteroids, but our study took place before evidence of beneficial effects of early short course of corticosteroids. Additional data suggest that corticosteroids benefit most to patients with CRP level higher than 20 mg/dL (15), striking our results. However, RECOVERY did not mandate positive RT-PCR to be included and randomized in the study (11% of the whole cohort). Interestingly, Hu *et al*.. found than headache was associated with intermittent negative SARS-CoV-2 RT-PCR status (16), highlighting our results about headache in the multivariate analysis.

Third, low rate of final diagnosis according to positive PCR testing is also a concern. Some protocols advocated positive PCR test to include patients (17) and we can hypothesize that physicians will be less prone to prescribe treatment to patients without positive PCR testing. Global sensitivity of RT-PCR is described as to 70% (18) but with major impact of duration between symptom onset and day of RT-PCR testing: between 38% of false negative at the day of symptom onset to 20% at day 8, and then false negative rate is increased once again(19). Similarly, Doll *et al* found than 100% of 19 patients with high probability of COVID-19 were subsequently tested as negative despite multiple testing (20). Ai *et al*., found than chest CT has a higher sensitivity for diagnosis of COVID-19 and may be considered as a primary tool for detection in epidemic area (21). Therefore, a strategy consisting to perform several testings to document virologic proof of COVID-19 can be debated.

Our study took place during the first epidemic wave in France and Belgium and was only dedicated to patients requiring hospitalization. It leads to high pre-test probability of COVID-19. We carefully selected hospitalized patients with several strong arguments for COVID-19 and final diagnosis of COVID-19 at hospital discharge.

Some limitations of our study must be highlighted. First, negative RT-PCR testing can be related to other disease. However, 45 (59.96%) of cases were also negative for other pathogen research during their hospital length stay and final diagnosis of COVID-19 was performed according to multimodal strategy including chest CT-scans in 88.75% of cases. Second, some issues could occur during technical sampling of RT-PCR but all RT-PCR were performed in hospitals with trained nurses and dedicated protocol to ensure high adherence to methods of

RT-PCR collection. Third, our sample size is limited, but we chose to restrain inclusion of patients with robust arguments for COVID-19 according to other diagnostic methods (especially chest CT-scans) with limited availability during epidemic wave in Europe. Last, we included patients from several centers with different RT-PCR detection kits. However, evidence suggest similar performance of available RT-PCR kits (22, 23).

## CONCLUSIONS

Patients with first negative RT-PCR testing for COVID-19 expected higher inflammatory markers, even at median duration of 6 days after symptom onset. Decision to perform or to withdraw special treatments such as corticosteroids for patients with COVID-19 cannot be done according only to virologic isolation of SARS-CoV-2. Multimodal strategy for diagnosis including radiological findings and clinical history is mandatory for each patient with suspected COVID-19.

## Data Availability

Data will be available upon request to corresponding author with protocol approved by IRB.

## ACKNOWLEDGEMENTS

We thank M. Rouaud, PharmD, for help during administrative process. We thank Mariana Ismael for Castor EDC (Amsterdam, The Netherlands) for technical support to design eCRF. We thank Pr C. Duclos MD, PhD, for her help in screening process in the University Hospital Center Avicenne (Bobigny, France).

